# A deep learning approach to identify seizure-prone and normal patients from their EEG records

**DOI:** 10.1101/2022.06.15.22276461

**Authors:** Sayantani Basu, Roy H. Campbell

## Abstract

Various learning models distinguish between an electroencephalogram (EEG) record of a normal patient and one having a seizure. In this paper, we propose a deep-learning based short-term memory (LSTM) model to identify whether an EEG record belongs to a seizure-prone patient with a non-seizure record or to a normal patient. The study builds on two datasets, namely the TUH Abnormal EEG Corpus (TUAB) and the TUH EEG Seizure Corpus (TUSZ) including the classified EEG records for seizure-prone and normal patients. We conducted experiments on both imbalanced and balanced datasets and show results using an LSTM model. We observed that the model performs consistently in both balanced and imbalanced cases using only 5 seconds of EEG data from the patient records. We show that our proposed LSTM model gives test accuracies up to 99.84% in case of 2-class classification between the non-seizure and normal classes and up to 98.87% in case of 3-class classification among non-seizure, seizure, and normal classes. This provides a basis for making improved temporal predictions about the occurrences of seizures.

## I. Introduction

Epilepsy is a neurological disorder characterized by the occurrence of seizures from the sudden firing of neurons. The electrical signals of the brain are recorded using electroencephalography and the corresponding record is known as an electroencephalogram (EEG). In case of patients with seizure disorders, the EEG shows indications of seizures which can be evaluated by medical professionals to provide a diagnosis for the patient and prescribe a treatment plan involving medications or surgical procedures. However, with the variations of seizure disorders, it may be difficult for medical professionals to constantly monitor the patient for seizures, especially in settings where EEG recordings are carried out over several hours. Moreover, it is tedious to view the recordings of such patients and manually forecast the onset of a seizure.

Motivation for our research concerns the TUH EEG Database. This dataset consists of scalp EEG recordings including patients with seizures. The files consist of recordings in .edf (European Data Format) with additional summary files of any seizures.

In this study, we consider looking at EEG records of seizure-prone and normal patients. The term ‘seizure-prone’ is indicative of the patient’s EEG having one or more seizures. It is important to note that even when patients have seizures, the frequency of seizure occurrence differs from patient to patient. It is also possible for a patient to not have a seizure during the time the EEG is being recorded. On the other hand, the term ‘normal’ is indicative that the patient does not have a seizure disorder at the time of the EEG recording nor during the recording. In this context, it is important to note that such patients may have had a history of seizures, which means they may have had seizures in the past or in their childhood.

Seizures are rare events in most cases, but their occurrence in a patient can be indicative of a long term condition that can be detected by our algorithms. Our results show:

1. Whether a patient is seizure-prone or normal based on a 5-second EEG sample.
2. They show that patients that have a tendency to have seizures are distinctively recognized by the LSTM model.
3. a foundation for a more sensitive and reliable test of whether a seizure might occur in the near future. This could benefit researchers who are trying to predict seizure events [1], [2].

Our results are important from a clinical point of view because they aid diagnosis and treatment for a useful scenario when clinicians are seeking to record an EEG of a patient; allowing then to record shorter EEGs or identifying potentially the sequences leading to an actual seizure.

The rest of this paper is structured as follows: Section II covers related work in this area, Section III explains our experimental setup, Section IV discusses our results, and Section V concludes the paper and discusses planned future work.

## II. Related Work

Several studies used deep learning approaches for the task of distinguishing between seizure and non-seizure records. Golmohammadi et al. [3] compared the performance of Long Short-Term Memory (LSTM) and Gated Recurrent Units (GRU) on the TUH EEG Seizure Corpus (TUSZ) for the task of classifying seizures and non-seizures. They evaluated the models using hybrid convolutional neural networks. They reported that convolutional LSTMs performed better and reported the best sensitivity of 30%. Shah et al. [4] have studied the performance of various channel selections for detection of seizures from the TUH EEG Seizure Corpus (TUSZ). They report the best results of 39% on using all 22 channels.

Many studies have modeled seizure prediction as a classification between interictal and preictal periods. Wei et al. [5] proposed a long-term recurrent convolutional network (LRCN) to detect seizures from data collected from the Xinjiang Medical University. They converted EEG recordings into images for applying their deep learning model. They obtained an accuracy of 93.4% with their proposed method. Cho et al. [6] proposed a model using various filtering algorithms on the CHB-MIT Scalp EEG Database [7], [8]. Their model using noise-assisted multivariate empirical mode decomposition (NA-MEMD) resulted in the highest accuracy of 83.17%. Another method on the CHB-MIT Scalp EEG Database proposed by Zhang and Parhi [9] used feature extraction as input to their support vector machine (SVM) model. They also tested their model on the intra-cranial EEG in the Freiburg Database [10]. They obtained sensitivities of 98.68% and 100% on the CHB-MIT Scalp EEG Database and the Freiburg Database respectively.

Some other seminal works in this area have been compared in Table I.

**TABLE I.** Other approaches related to studies on seizures from EEG data

| Paper | Dataset | Method(s) | Notes | Performance |
| --- | --- | --- | --- | --- |
| [11] | TUH Abnormal EEG Corpus | 1D CNN | Considered temporal to occipital channels | 20.66% error rate |
| [12] | TUH EEG Corpus | SVM, kSVM, RF, SCNN, DCNN | Studied both feature and deep learning based methods | 73% accuracy |
| [13] | TUH Abnormal EEG Corpus (TUAB),<br>TUH EEG Artifact Corpus (TUAR),<br>TUH EEG Seizure Corpus (TUSZ) | InstaGATs,<br>LSTM+attention,<br>CNN+attention | LSTM methods had highest accuracy for TUAB and TUAR, CNN methods performed better for TUSZ | Accuracies of 79.48% (TUAB), 83.15% (TUAR), 96.98 % (TUSZ) |
| [2] | CHB-MIT Scalp EEG Database | LSTM | Extracted set of features for preictal-interictal classification | 100% of seizures detected, 0 false alarms in 17/24 patients |
| [14] | TUH EEG Corpus (TUH-EEG) | HMMs,<br>HMM/deep learning | Used 22 channels for experiments | sensitivity>90%, specificity<5% |
| [15] | TUH EEG Seizure Corpus | Attention-gated U-nets | Combination of U-nets and LSTM for prediction on 16 channels | sensitivity<25% |
| [16] | CHB-MIT Scalp EEG Database | Deep neural network | Discussed both inter and intra patient detection | sensitivity 85% |

We would like to summarize our technical contributions as follows:

i. We propose a long short-term memory (LSTM) [17] based deep learning model that can classify EEGs as non-seizure, seizure, and normal (seizure-free). We ran an additional set of experiments to determine whether the samples are non-seizure or normal.
ii. We have used samples from TUSZ and TUAB to train and test our proposed model. We believe our study is one of the first attempts involving a classification model on multiple types of corpora.

## III. Experimental Setup

### A. Datasets

For this project, we used two corpora from Temple University Hospital (TUH) Dataset [18] – the TUH Abnormal EEG Corpus (TUAB) and the TUH EEG Seizure Corpus (TUSZ). For TUAB, we used version v2.0.0 and for TUSZ, we used version v1.5.2. For the labels of seizure and non-seizure, we used the reference file for the train set of TUSZ. For the seizure-prone patients, we used the label of ‘seiz’ to indicate a seizure period, and ‘bckg’ to indicate a non-seizure period from TUSZ. For the normal patients, we used data from the ‘normal’ sub-folder of TUAB. The final data considered in this study consists of the following labels:

1. *non-seizure*: Patients who are clinically diagnosed with seizures where the EEG sample retrieved does not contain seizures.
2. *seizure*: Patients who are clinically diagnosed with seizures where the EEG sample retrieved contains seizures.
3. *normal*: Patients whose EEG records do not show any clinical abnormalities.

### B. Preprocessing

Each EEG recording was divided into overlapping segments of 5 seconds each. The data was extracted from the .edf files using the Python MNE package. For this paper, we chose to analyze records sampled at 256Hz with 26 common channels. These channels are: FP1-REF, FP2-REF, F3-REF, F4-REF, C3-REF, C4-REF, P3-REF, P4-REF, O1-REF, O2-REF, F7-REF, F8-REF, T3-REF, T4-REF, T5-REF, T6-REF, T1-REF, T2-REF, FZ-REF, CZ-REF, PZ-REF, EKG1-REF, C3P-REF, C4P-REF, SP1-REF, and SP2-REF. Channel selection for our experiments choose record availability for the set of *n* channels. The selection of *n* as 26 was also challenging as we had to select sufficient records across the two corpora. All the data was normalized using StandardScaler from scikit-learn [19] for the samples, and normalized using z-score for the channels. Prior to the model training, the 5-second segments were always shuffled along with their corresponding labels.

### C. Performance Metrics

We used accuracy as the metric to determine performance. Our main focus is the test accuracy as it helps understand the true model performance and generalization on the test dataset. However, we also report the confusion matrices for all the experiments to show the model performance across all classes.

### D. Training and Testing

All our experiments were run on gpux1 of the HAL cluster [20]. This gives results on 80% of training samples and 20% of testing samples. All experiments were coded using Python3 in Keras [21] using Tensorflow [22] as the backend.

## IV. Results and Discussion

We trained the LSTM model on 80% of the samples and report test results on 20% of the samples.

The various experiments performed are discussed below:

- Imbalanced 2-class: Based on our constraints for the number of channels, the sampling frequency, and the metadata availability, we had 289 patients from TUSZ and TUAB with 139,520 train and 34,816 test EEG samples in total. In this experiment, we label samples only as non-seizure or normal (seizure-free). We leave out EEG samples labeled as seizures.
- Imbalanced 3-class: Based on our constraints for the number of channels, the sampling frequency, and the metadata availability, we had 289 patients from TUSZ and TUAB with 151,552 train and 37,888 EEG samples in total. In this experiment, we label samples as non-seizure, seizure, or normal (seizure-free).
- Balanced 2-class: The first set of experiments had a larger number of patients from TUSZ compared to TUAB. To show the consistency in our results, we show results on a balanced dataset with equal proportions of patients from TUSZ and TUAB. Based on our constraints for the number of channels, the sampling frequency, and the metadata availability, we had 26 patients from TUSZ and 26 patients from TUAB with 40,704 train and 9,984 EEG samples in total. In this experiment, we label samples only as non-seizure or normal (seizure-free). We leave out EEG samples labeled as seizures.
- Balanced 3-class: Similar to the balanced 2-class experiment, in order to show the consistency in our results, we show our results on a balanced dataset with equal proportions of patients from TUSZ and TUAB. Based on our constraints for the number of channels, the sampling frequency, and the metadata availability, we had 26 patients from TUSZ and 26 patients from TUAB with 41,472 train and 10,240 EEG samples in total. In this experiment, we label samples as non-seizure, seizure, or normal (seizure-free).

For each of the above experiments, we retained the same network architecture for the long short-term memory (LSTM) model, except for the final dense layer which had to be changed depending on the number of classes considered for classification (2 or 3). Based on our experiments, we obtained consistent performance using 3 LSTM layers with 128 units each, interleaved with dense layers containing 25 units each and ReLU activation. The final dense layer has 2 or 3 units depending on the number of classes and softmax activation. The stacked LSTM architecture was selected based on empirical performance.

We used Adam for optimization with 0.001 learning rate. In each fold, we trained for 25 epochs with a batch size of 256. All classes were weighted based on the ratio of the total number of samples and number of samples for each class. We used the MirroredStrategy from Tensorflow for distribution on the GPU and adjusted our learning rate and batch size accordingly.

Performance metrics are reported in terms of accuracy on the test data.

As shown in Table II, we show our results are consistent in both the balanced and imbalanced cases. This shows that an imbalance in the number of records from the TUAB and TUSZ corpora does not significantly affect model performance. This is important in a clinical context because there may be different ratios of seizure-prone and normal patients available as training data. While the model would be usable in the case of imbalanced records in a clinical context, based on the results, it is preferable to use a balanced dataset if possible. The test accuracies for 2-class classification for both experiments showed higher accuracy compared to the test accuracies for 3-class classification. The confusion matrices in Table III, Table IV, Table V, and Table VI provide further insight on the classwise performance of the LSTM model for the imbalanced 2-class, imbalanced 3-class, balanced 2-class, and balanced 3-class experiments respectively. In all confusion matrices, *t* indicates the true labels and *p* indicates the predicted labels.

**TABLE II.** Performance of proposed LSTM model

| Experiment | Train | Test | Accuracy |
| --- | --- | --- | --- |
| Imbalanced 2-class | 139520 | 34816 | 99.84% |
| Imbalanced 3-class | 151552 | 37888 | 98.87% |
| Balanced 2-class | 40704 | 9984 | 97.89% |
| Balanced 3-class | 41472 | 10240 | 98.53% |

**TABLE III.** Confusion Matrix for Imbalanced 2-class

| Label | Non-seizure (t) | Normal (t) |
| --- | --- | --- |
| Non-seizure (p) | 27321 | 26 |
| Normal (p) | 28 | 7441 |

**TABLE IV.** Confusion Matrix for Imbalanced 3-class

| Label | Non-seizure (t) | Seizure (t) | Normal (t) |
| --- | --- | --- | --- |
| Non-seizure (p) | 27297 | 170 | 3 |
| Seizure (p) | 209 | 2715 | 0 |
| Normal (p) | 43 | 4 | 7447 |

**TABLE V.** Confusion Matrix for Balanced 2-class

| Label | Non-seizure (t) | Normal (t) |
| --- | --- | --- |
| Non-seizure (p) | 2534 | 167 |
| Normal (p) | 44 | 7239 |

**TABLE VI.** Confusion Matrix for Balanced 3-class

| Label | Non-seizure (t) | Seizure (t) | Normal (t) |
| --- | --- | --- | --- |
| Non-seizure (p) | 2690 | 18 | 15 |
| Seizure (p) | 41 | 142 | 2 |
| Normal (p) | 75 | 0 | 7257 |

## V. Conclusion and Future Work

In this paper, we studied two datasets, namely the TUH Abnormal EEG Corpus (TUAB) and the TUH EEG Seizure Corpus (TUSZ) [18] and classified EEG samples from records of seizure-prone and normal patients. We conducted experiments on both imbalanced and balanced datasets and show results with a deep learning based long short-term memory (LSTM) model. We observed that the performance is consistent for both balanced and imbalanced datasets based on only 5 second samples from the EEG records. We show that our proposed LSTM model gives test accuracies up to 99.84% in case of 2-class classification between the non-seizure and normal classes and up to 98.87% in case of 3-class classification among non-seizure, seizure, and normal classes.

In the future, we plan to extend this work by using other deep learning models for performing classification tasks on the TUH EEG database.

## Data Availability

All datasets are available online at: https://isip.piconepress.com/projects/tuh_eeg/html/downloads.shtml

https://github.com/sayantanibasu/eeg-seizure-normal

## VI. Code

Code for this paper is available at this link: https://github.com/sayantanibasu/eeg-seizure-normal.

## Acknowledgment

This work utilizes resources supported by the National Science Foundation’s Major Research Instrumentation program, grant #1725729, as well as the University of Illinois at Urbana-Champaign.

